# Back to the Basics: An Alternative Approach to ABG Interpretation

**DOI:** 10.1101/2023.08.21.23294231

**Authors:** Emily Nguyen, Chase Turpin, Andres deLuna

## Abstract

Medical education has promoted the rapid interpretation of the arterial blood gas (ABG) as a crucial skill in the management of critically ill patients. The first step of many algorithms focuses on using log-linear estimates to determine the internal consistency of ABG values. However, because of modern computing advances, these estimates are no longer necessary. We demonstrate an alternative method for ascertaining internal consistency of ABG values based on direct calculation derived directly from Henderson-Hasselbach and demonstrate its efficiency and accuracy.

Deidentified ABG values were collected from electronic medical records for the month of September 2022. Estimated hydrogen concentration was calculated by log linear estimation and by the proposed method of direct calculation. Time of computation, a measure of efficiency, was also recorded.

Of the 1008 ABGs used for analysis, 185 (18.4%) had estimated hydrogen concentrations that were greater than a 5% threshold according to the guidelines method, suggesting potential internal inconsistency. However, only 13 (1.3%) had estimated hydrogen concentrations greater than 5% by direct calculation. Analysis was also performed at a 10% threshold. On further analysis, 4 ABGs that were identified by the guidelines method as being internally inconsistent, but not identified by the direct calculation method, were found to have pH <7.0, outside the range provided for the current guidelines for log-linear estimation, and were thus incorrectly identified by the guidelines method as being internally inconsistent. The time of computation for direct calculation was approximately 8-fold faster, although the total time of calculation for this dataset was minimal.

This study demonstrates a direct calculation method as an alternative approach to traditional guidelines to assess internal ABG value consistency, the first step of ABG interpretation. We demonstrate that direct calculation is more accurate, identifying less potentially internally inconsistent values, notably at extremes in pH. In addition, the direct calculation algorithm is 8-fold more efficient than guidelines-based algorithm, although actual processing times were miniscule for the data set regardless of the method used. Furthermore, with modern calculators and computers, the direct calculation method is easier to understand and implement than the guideline based approach. Therefore, based on the results of this study, we propose the use of direct calculation as an alternative to log-linear based estimation for the assessment of internal consistency of ABGs.

## Introduction

Medical education has promoted the rapid interpretation of the arterial blood gas (ABG) as a crucial skill in the management of critically ill patients. The first step of many algorithms focuses on determining the internal consistency of ABG values based on the Henderson-Hasselbach equation. The current method for these algorithms was based on log-linear estimation rather than direct calculation, as proposed by a landmark study (1), which demonstrates the linear relationship of pH and estimated H+ at physiologic pH values. The lack of an efficient means for logarithmic calculations was the impetus for the use of this method, with slide-rules being often used at the time to estimate logarithms for calculations. However, these log-linear estimates are no longer necessary because almost any modern calculator can perform logarithmic calculations. Therefore, the current study demonstrates an alternative method for ascertaining internal consistency of ABG values. Specifically, the current study aims to show the utility of an alternative method for ascertaining internal consistency of ABG values based on direct calculation through the use of real-world and simulated data sets.

## Methods

The standard Henderson-Hasselbach equation is used to demonstrate the derivation of a method for direct calculation of H+ concentration, [H+], from pCO2 and HCO3 (Figure 1). This H+ can be directly compared to [H+] as defined by pH. The validity of the ABG can be assessed by the degree of discrepancy between [H+] derived from pCO2 and HCO3 and the [H+] derived from pH. Deidentified ABG values were collected from electronic medical records for the month of September 2022 from a single institution. Because the aim of the study was to demonstrate a method for ABG interpretation, detailed patient information was deemed unnecessary. The Institutional Review Board (IRB) of Santa Clara Valley Medical Center declared the to be exempt from further IRB review on September 12, 2022, under exemption 45 CFR 46.104(d)(2)(i).

**Figure 1.**
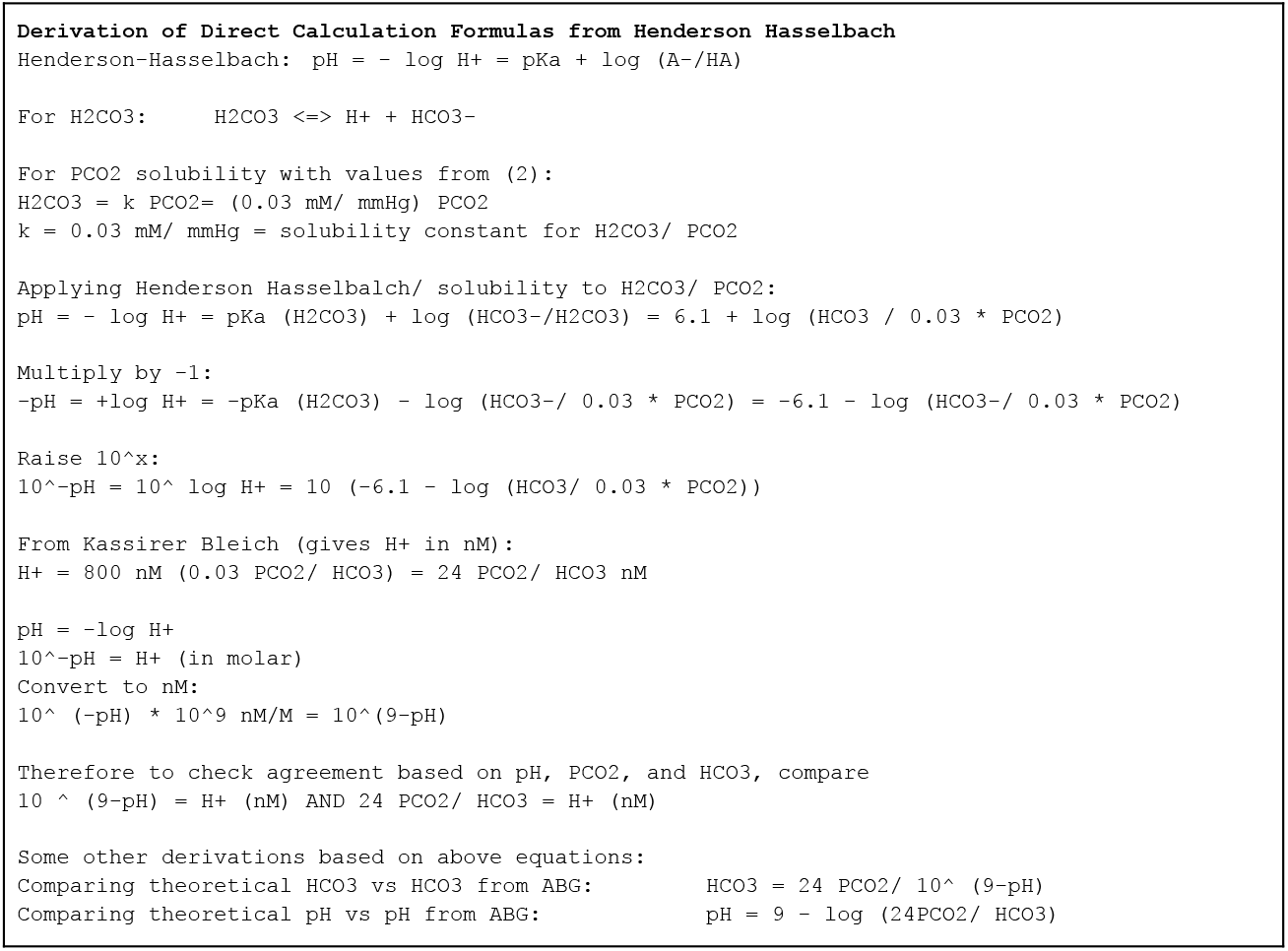

A total of 3381 ABG components (pH, pCO2, HCO3) were collected and merged. Duplicate component values and ABGs with missing data were deleted. A total of 1008 complete ABGs were obtained for analysis. Estimated hydrogen concentration was calculated by log linear estimation and by the proposed method of direct calculation as described above. Time of computation, a surrogate measure of efficiency, was also calculated. All statistical computations were performed using the R statistical software package.

## Results

A total of 1008 complete ABGs were used for analysis. For the complete set of ABGs used, the values were (mean, SD): pH(7.39, 0.10), pCO2 (41, 13) and HCO3 (24, 6). Of the 1008 ABGs, 185 (18.4%) had estimated hydrogen concentrations that were greater than a 5% threshold according to the guidelines method, suggesting potential internal inconsistency.

However, only 13 (1.3%) had estimated hydrogen concentrations greater than 5% by direct calculation. Analysis was also performed at a 10% threshold, which was thought to be more clinically relevant. At the 10% threshold, 16 (1.6%) had estimated hydrogen concentrations that were greater than 10% according to log-linear estimation, whereas 12 (1.2%) had estimated hydrogen concentrations that were greater than 10% according to direct calculation (Table 1).

**Table 1.**

|  | Log Linear Estimation | Direct Calculation |
| --- | --- | --- |
| ABGs lacking internal consistency at the 5% threshold | 185 (18.4%) | 13 (1.3%) |
| ABGs lacking internal consistency at the 10% threshold | 16 (1.6%) | 12 (1.2%) |

On further analysis, the 4 ABGs that were identified by the guidelines method as being internally inconsistent, but not identified by the direct calculation method, were found to have pH <7.0, outside the range provided for current guidelines (3) for log-linear estimation. These ABGs were thus incorrectly identified by the guidelines method as being internally inconsistent (Table 2).

**Table 2.**
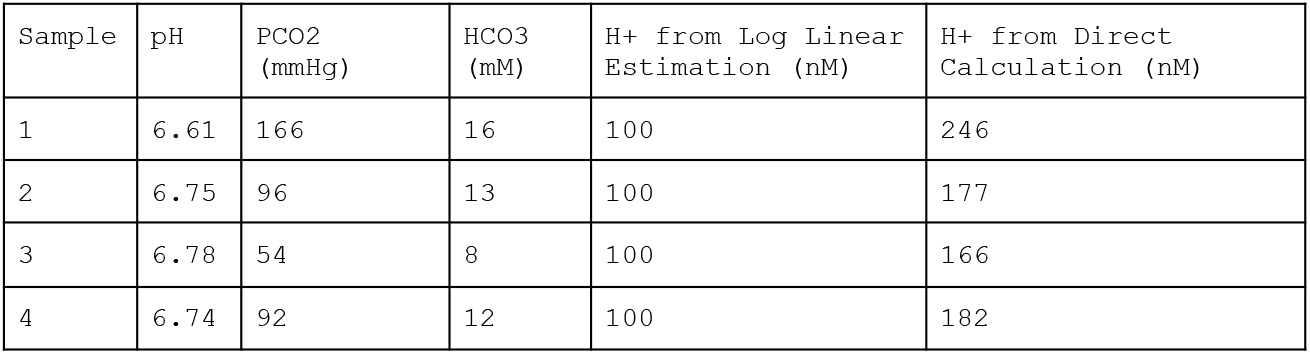

The time of computation for the log-linear method was 0.0493 seconds, whereas the time of computation for direct calculation was 0.0061 seconds. Thus, direct calculation was approximately 8-fold faster, although the total time of calculation for this dataset was minimal.

## Discussion

This study proposes a direct calculation method as an alternative approach to log-linear estimation to assess internal ABG value consistency, the first step of ABG interpretation.

We demonstrate that direct calculation identifies less potentially internally inconsistent values, especially at extremes in pH. The identification of less internally inconsistent values results in decreased need to repeat ABGs before making clinical decisions. Thus, not only are resources saved if less ABGs are repeated, but also clinical decisions can be made faster, as one does not have to wait for the repeat ABG. Perhaps more important, the current guidelines method does not work for extremes in pH, i.e. pH<7.0 or pH>7.65. These patients are more ill and thus would benefit from more timely clinical decisions, which should not be impeded by inaccurate labeling of an ABG as being potentially internally inconsistent because estimates for a given pH are not available.

In addition, the direct calculation algorithm was 8-fold faster than guidelines-based algorithm, although actual processing times were miniscule for this data set regardless of the method used. The code for the guidelines-based log linear estimation method involves the assignment of different [H+] values to a given pH, based on the category of the pH value. However, the code for direct calculation is a single line of code for calculation based on the pH. This efficiency in coding reduces processing times. For a small data set such as this one (∼ 1000 samples), the processing time is miniscule. However, for much larger data sets (eg 10^9 samples), the processing time becomes more important.

Furthermore, with modern calculators and computers, direct calculation is easier to understand and implement than log-linear estimation. The use of log-linear estimation requires more rote memorization of [H+] values than the direct calculation method. If one is to understand the derivation of these values, one must also have a grasp of log-linear estimation and its derivation (2). In contrast, direct calculation requires only an elementary understanding of logarithms and exponents for calculations. Thus, direct calculation may be more easily grasped to students learning ABG interpretation for the first time.

Therefore, based on the results of this study, we propose the use of direct calculation as an alternative to log-linear based estimation for the assessment of internal consistency of ABGs.

## Data Availability

All data produced in the present study are available upon reasonable request to the authors

## Notes

### Competing Interest Statement

The authors have declared no competing interest.

### Funding Statement

This study did not receive any funding

### Author Declarations

The Institutional Review Board (IRB) of Santa Clara Valley Medical Center waived ethical approval of this work.

## References

1. Kassirer JP, Bleich HL. Rapid Estimation of Plasma Carbon Dioxide Tension from pH and Total Carbon Dioxide Content. N Engl J Med. 1965 May 20;272:1067–8. doi: 10.1056/NEJM196505202722007. PMID: 14281544.

2. Rose BD Post TW. Clinical Physiology of Acid Base and Electrolyte Disorders. 6. ed. New York: McGraw-Hill; 2017.

3. Kaufman, David. Interpretation of Arterial Blood Gases. American Thoracic Society. https://www.thoracic.org/professionals/clinical-resources/critical-care/clinical-education/abgs.php

